# A distributed learning strategy improves performance and retention of skills in neonatal resuscitation: A simulation-based randomized controlled trial

**DOI:** 10.1101/2020.10.28.20221473

**Authors:** Pratheeban Nambyiah, Sylvain Boet, Gregory Moore, Riley Boyle, Deborah Aylward, Andre Jakubow, Sandy Lam, Karim Abdulla, M. Dylan Bould

## Abstract

Skill retention after neonatal resuscitation training is poor. A distributed learning strategy – where learning is spread over multiple sessions – can improve retention of declarative memory (facts & knowledge). Session timings are critical – maximal retention occurs when a refresher session is scheduled at 10-30% of the time between initial training and test. We hypothesized this also holds true for neonatal resuscitation, a complex skill set requiring both declarative and procedural memory. We conducted a prospective, single-blinded randomized-controlled trial. University of Ottawa residents were recruited to training in neonatal resuscitation, with a high-fidelity simulated pre-test, immediate post-tests, and a retention test at 4 months. After training, they were randomized to either a refresher session at 3 weeks (18% of interval) or at 2 months (50%). Technical and non-technical skills were scored using validated checklists, knowledge with standardized questions. There was no difference between groups prior to the retention test. The early refresher group demonstrated significantly improved technical (mean ± 95% CI: 22.4 ± 1.3 v 18.2 ± 2.5, p = 0.02) and non-technical (31.0 ± 0.9 v 25.6 ± 3.1, p = 0.03) skill scores in the retention post-test compared to the late group. No difference was seen with knowledge scores. We conclude that both technical and non-technical aspects of neonatal resuscitation performance can benefit from an early refresher session. Session timings are critical and should be tailored to the desired length of skill retention. Findings may be generalizable to other interventions that depend on mixed types of memory.

## Introduction

Ten percent of babies born in the high-income world require medical intervention at birth^1^. The Neonatal Resuscitation Program® (NRP®) developed by the American Academy of Pediatrics (AAP) reflects international consensus on best practice^2^. Similar courses for neonatal, child and adult resuscitation are run elsewhere. They teach knowledge and skills, improve performance in simulated settings^3-6^, and contribute to improved patient outcomes^6-8^. Unfortunately, attrition of these skills occurs over time^5,9-11^. The consensus document from the International Liaison Committee on Resuscitation (ILCOR) advocates educational strategies which improve retention^12^.

There is consensus that training followed by long intervals without formal training is undesirable^2,12^. Recent studies favor approaches which employ e-learning, modularity, and simulation^13-15^. However, heterogeneity is an obstacle to evaluating and standardizing practice, and to achieving value for money. One fundamental problem is the lack of quantitative theoretical frameworks on which to base practical training structures. Cognitive psychology studies have established that distributed learning, where a fixed amount of study time is broken into intervals (e.g. one hour of training for five days) rather than all at once (e.g. five hours of training in a single day), benefits long-term retention of knowledge^16-18^. Most of these studies investigated learning corresponding to declarative memory – knowledge that can be consciously recalled. In medicine, procedural memory is also important, but the evidence for distributed learning is mixed – for example, microvascular anastomosis was shown to benefit from a distributed learning strategy^19^, whereas no difference could be shown with bronchoscopy^20^.

Learning during the NRP involves both declarative and procedural memory, executed under time-critical situations when rapid memory recall is imperative to patient survival. In an emergency context, the retention of procedural memory is poorer than declarative memory^21,22^. The evidence in favor of distributed learning in resuscitation skills is not convincing. Distributed learning can be organised in the form of a refresher course after initial training. Established resuscitation training programs offer renewal courses after intervals of a year or more, but they do not provide a refresher session soon after initial certification. The effect of a resuscitation refresher has been studied^9,23^, but most investigators have been unable to show benefit. These studies provided a refresher several months after initial teaching and tested for retention relatively soon after. Evidence from the field of cognitive psychology suggests that the interval between original training session and refresher (the inter-study interval, ISI), and the interval between original session and retention test (the retention interval, RI), are important determinants of success (Fig. 1)^24,25^. In a review of literature, Cepeda et al. note ‘the optimal ISI increases as the duration over which information needs to be retained increases’^24^; i.e. if one wants to retain information for a longer period, one must increase the ISI. In a subsequent review, Rohrer and Pashler (2007)^25^ conclude that ‘the optimal ISI [lies] at a value of 10 to 30% of the RI’. This may explain why previous researchers looking at distributed resuscitation learning failed to find an effect – their ISI:RI ratios were greater than 50%^9,23^.

**Fig. 1.**
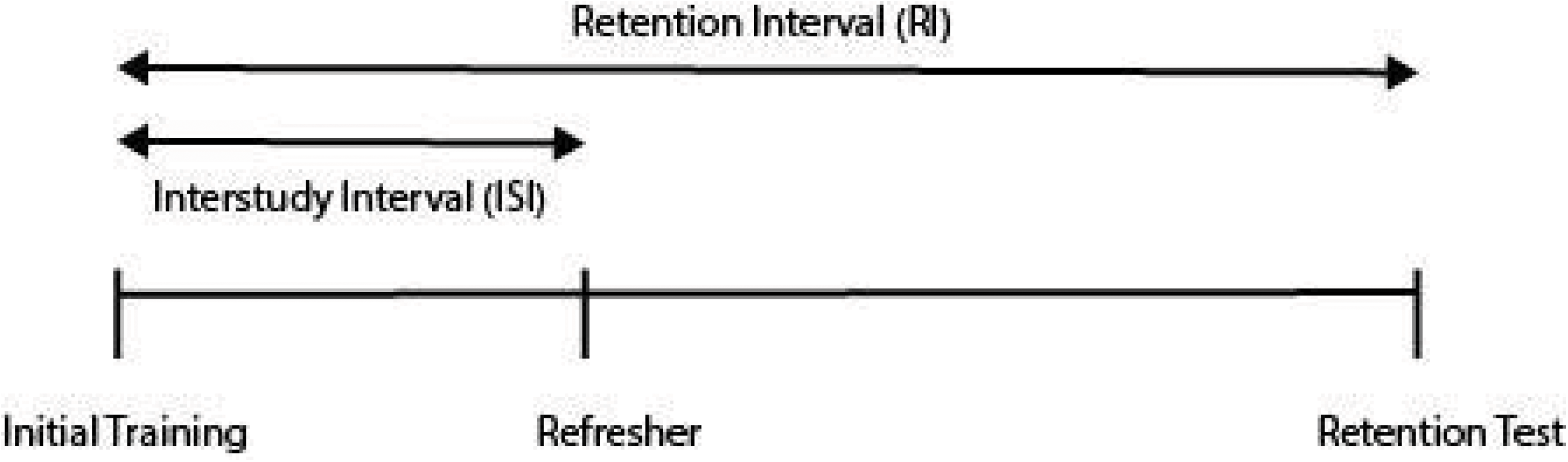
A distributed learning program, with initial training, refresher and retention test. Definitions of retention interval and inter-study interval are given.

### Hypothesis

This study aims to evaluate whether an evidence-based distributed learning strategy can improve retention of performance and knowledge in neonatal resuscitation. We hypothesize that a refresher session at three weeks after initial training is more effective than a refresher at two months, for the retention of performance and knowledge at four months. In the early refresher group, ISI will be 18% of RI – within the optimal range identified above. In the late refresher group, ISI will be 50% of RI.

## Methods

Ethical approval was secured from the Ottawa Health Science Network Research Ethics Board (Protocol #20120849-01H). We obtained written informed consent from each participant.

Participants were University of Ottawa residents in the departments of anesthesiology, family medicine and emergency medicine. They were selected after a call for interested volunteers approved by the programme leads. A questionnaire was administered to collect demographic data, including specialty, postgraduate year, and experience in neonatal resuscitation management. Residents were excluded if they were unable to commit to all phases of the study, or if they were NRP instructors.

This was a prospective, single-blinded randomized controlled trial (Fig. 2). Prior pilot data was collected using volunteers, allowing us to ‘dry run’ scenarios. Videos collected from pilot sessions were used for rater training and calibration. Participants underwent a two-hour training session in neonatal resuscitation as per NRP 6^th^ edition guidelines (G.M. and D.A., NRP instructors). Before the session began on day 1, they completed a multiple-choice knowledge test (‘written pre-test’, with questions taken from the NRP bank) and performed a simulated standardized NRP scenario (‘simulation pre-test’). Following training, they took the same written and simulation tests (‘immediate post-tests’). Structured content debriefing using the NRP checklist was given after the simulation post-test to maximise educational benefit. Fifteen minutes were allocated for the scenario and fifteen minutes for debrief. All participants were then randomized to an early refresher (intervention) or late refresher group (control). Block randomization was done using sealed envelopes and was stratified by specialty and years of postgraduate experience. The intervention group received an early refresher session at three weeks post-training. The refresher consisted of the same simulated resuscitation scenario, but this time *in situ* at the participant’s usual place of work, followed by structured content debriefing. Again, fifteen minutes each were allocated to scenario and debriefing. This structure made use of two evidence-based strategies that have been shown to aid retention^26^. Firstly, the ‘testing effect’ (the fact that actively recalling information from memory strengthens later retention); secondly, timely feedback. The control group received a late refresher session at two months post-training. From the work by Kaczorowski and colleagues, a refresher at this time appears to be ineffective, and therefore an appropriate control^9^. All participants underwent retention post-tests at 4 months. This consisted of the same simulation scenario and written test. A questionnaire was administered after the retention tests to collect information on clinical neonatal resuscitation experience gained since initial training.

**Fig. 2.**
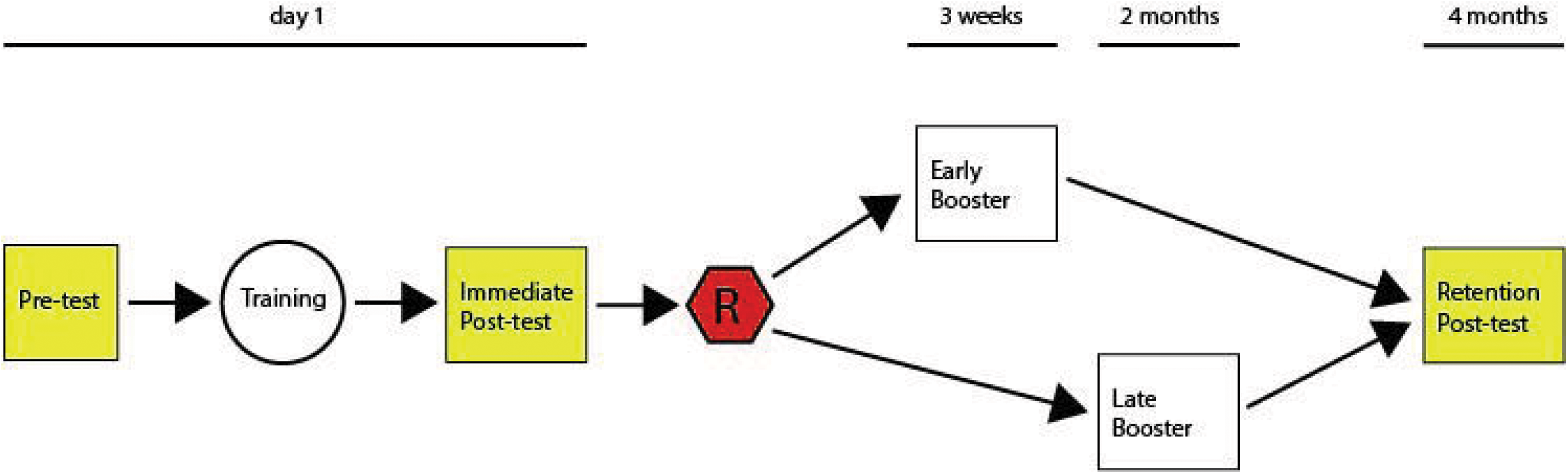
Trial structure. R = randomization.

Participants were asked to perform the resuscitation scenario in a simulated environment with a Newborn HAL S3010 simulator (Gaumard Scientific, Miami, FL.). In this scenario, a term neonate is delivered by caesarean section. The subject is told the child is ‘flat’, and asked to assess and initiate appropriate management. Simulator settings were: (i) apnoea; (ii) HR 50; (iii) SpO_2_ 85% at delivery, deteriorating to 65% over 1min; (iv) cyanosis; (v) airway obstruction until appropriate manoeuvres performed. The performance was recorded for later analysis. All appropriate equipment was available as described in the AAP and Canadian Paediatric Society (CPS)’s NRP 6^th^ edition^27^. Two confederates were available: one to assist with NRP tasks, the other to find equipment and follow instructions.

The standardized, previously validated NRP Basic Megacode Assessment checklist (Canadian adaptation)^28^ (Appendix 1) was used to assess simulation performance, with a modification to reflect the fact that participants were not given the opportunity to demonstrate post-resuscitation care measures. We assessed non-technical skills using the Ottawa Global Rating Scale (GRS)^29^, a validated 7-point Likert scale which assesses performance in leadership, problem solving, situational awareness, resource utilisation and communication.

Two expert blinded raters (G.M & A.J) independently assessed videos. An inter-rater correlation coefficient was calculated on the experimental data-set. Knowledge was assessed with thirty questions from the NRP multiple-choice question bank, administered immediately after each scenario (Appendix 2). These were scored according to standard NRP criteria, with assessors blinded to study group.

The primary outcome measure was NRP Basic Megacode checklist score. Secondary outcome measures were: (i) Knowledge Scores on written tests, and (ii) Ottawa GRS scores.

Data was analyzed using SPSS 21.0 (SPSS Inc., Chicago, IL). The inter-rater reliability was measured using single-measures intraclass correlation coefficients (for total scores). NRP Basic Megacode checklist performance at pre-test and immediate post-test was assessed using analysis of variance (ANOVA). The performance score was treated as the dependent variable. The independent variables were the type of refresher (early vs. late) as the between-subjects variable, and the test phase (pre-test vs. immediate post-test) as the within-subjects variable. At retention post-test, NRP Basic Megacode checklist performance was assessed using one-way analysis of co-variance (ANCOVA). The retention performance score was treated as the dependent variable with the type of refresher (early vs. late) as fixed factor and immediate-post test score as the covariate. Written test and GRS scores were similarly analyzed using ANOVA and ANCOVA.

Sample size was calculated as follows. Previously, Bould and colleagues used the neonatal resuscitation performance checklist developed by Lockyear and colleagues^30,31^, and found that the mean score was 18/30 with a standard deviation of 4.16. Assuming an F test, an alpha error of 0.05, and a beta error of 0.20, we will have 80% power for detecting an increase to 22/30 on this checklist (corresponding to 2 additional items done correctly that were previously not done) with 19 subjects in each group at retention test^32^.

## Results

Eighteen participants were randomized. Seventeen completed all stages. One participant was unable to return for the refresher. Fig. 3 shows participant flow through the study. Table 1 summarizes the characteristics of participants who completed the program. Of note, we were unable to reach the target sample size due to difficulties with recruitment and drop outs after a prolonged recruitment phase. For these logistical reasons, we took the decision to terminate recruitment, and analyze findings. There was no significant difference in out-of-study exposure to neonatal resuscitation after randomization (Fisher exact test value = 1).

**Table 1.** Characteristics of study participants.

| Characteristic | Early Refresher Group | Late Refresher Group |
| --- | --- | --- |
| **Gender** |  |  |
| F | 5 | 7 |
| M | 2 | 3 |
| **Age (mean (sd))** | 30.9 (4.5) | 32.6 (5.1) |
| **Specialty** |  |  |
| Anesthesiology | 6 | 6 |
| Emergency Medicine | 1 | 2 |
| Family Medicine | 0 | 2 |
| **Previous NRP training** |  |  |
| Yes | 4 | 2 |
| No | 3 | 8 |
| **Year of Specialty Training** |  |  |
| 1 | 1 | 1 |
| 2 | 1 | 4 |
| 3 | 0 | 1 |
| 4 | 2 | 1 |
| 5 | 3 | 3 |
| **Neonatal resuscitation experience after recruitment** |  |  |
| Yes | 3 | 4 |
| No | 4 | 6 |

**Fig. 3.**
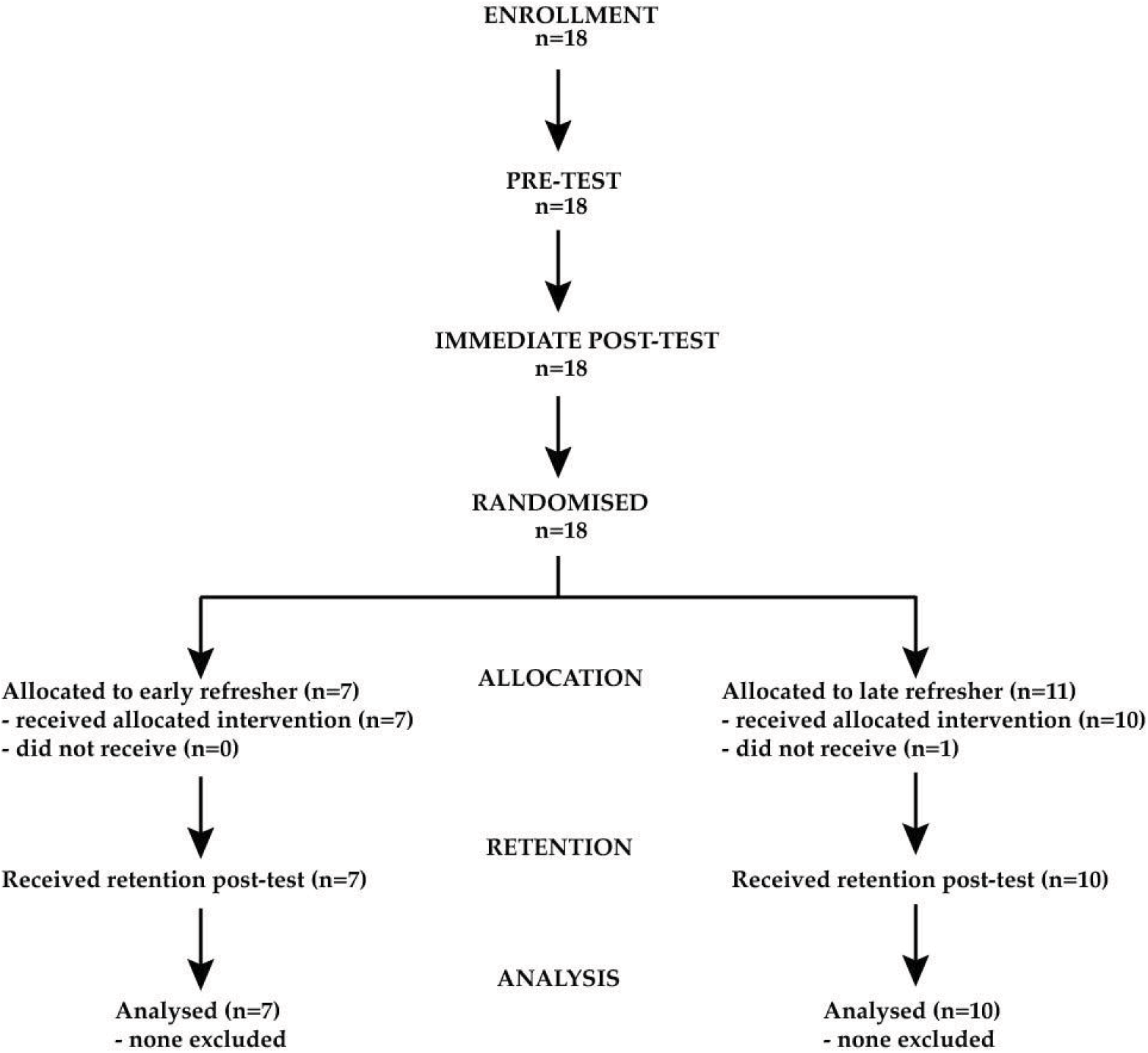
Patient flow chart.

Inter-rater reliability as measured using intra-class correlation coefficients (ICC). ICC for the NRP checklist was 0.74; for the Ottawa GRS it was 0.45. Following guidelines suggested by Landis and Koch, these indicate substantial and moderate agreement respectively^33^. Reduced inter-rater reliability for non-technical assessment has been seen in other studies, and our ICCs are in line with figures reported elsewhere for similar measures^13,34^.

Table 2 shows NRP checklist, Ottawa GRS, and knowledge test scores for early and late refresher groups at pre-test, immediate post-test, and retention post-test stages. For pre-test and immediate post-test, ANOVA shows no difference between groups for any outcome measure, suggesting that the groups were equally distributed prior to randomization. Both groups improved their scores from pre-test to immediate post-test, but ANCOVA shows no significant difference between the groups in the degree of this improvement (NRP checklist F(1,14) = 2.93, p = 0.11; Ottawa GRS F(1,14) = 3.07, p = 0.10; Knowledge test F(1,14) = 3.19, p = 0.10).

**Table 2.** NRP Checklist, Ottawa GRS and knowledge test scores at each experimental stage.

| Experimental Stage | Early Refresher | Late Refresher | ANOVA/ANCOVA |  |
| --- | --- | --- | --- | --- |
|  | mean (sd) | mean (sd) | F | p |
| **Pre-test (range)** |  |  |  |  |
| NRP Checklist (0-30) | 14.5 (3.6) | 11.8 (3.6) | F(1,15) = 2.624 | 0.13 |
| Ottawa GRS (6-42) | 21.9 (4.1) | 19.7 (4.4.) | F(1,15) = 1.124 | 0.31 |
| Knowledge Test (0-30) | 21.4 (2.4) | 21.5 (4.0) | F(1,15) = 0.002 | 0.97 |
| **Immediate Post-test (range)** |  |  |  |  |
| NRP Checklist (0-30) | 22.1 (3.7) | 20.0 (2.2) | F(1,15) = 2.299 | 0.15 |
| Ottawa GRS (6-42) | 31.1 (3.3.) | 28.4 (3.2) | F(1,15) = 2.927 | 0.11 |
| Knowledge Test (0-30) | 28.4 (1.0) | 27.3 (1.5) | F(1,15) = 3.047 | 0.10 |
| **Retention Post-test (range)** |  |  |  |  |
| NRP Checklist (0-30) | 22.4 (1.7) | 18.2 (4.0) | F(1,14) = 6.602 | 0.02 |
| Ottawa GRS (6-42) | 31.0 (1.2) | 25.6 (5.0) | F(1,14) = 5.564 | 0.03 |
| Knowledge Test (0-30) | 26.7 (1.5) | 26.5 (2.4) | F(1,14) = 0.009 | 0.93 |

Fig. 4 presents boxplots of outcome scores at each experimental stage. Both early and late refresher groups improved at retention post-test compared to pre-test scores. However, performance in the early refresher group was significantly better than the late refresher group for the primary outcome measure, NRP checklist scores. The early refresher group also demonstrated superior Ottawa GRS scores. No significant difference in retention was seen with written knowledge scores.

**Fig. 4.**
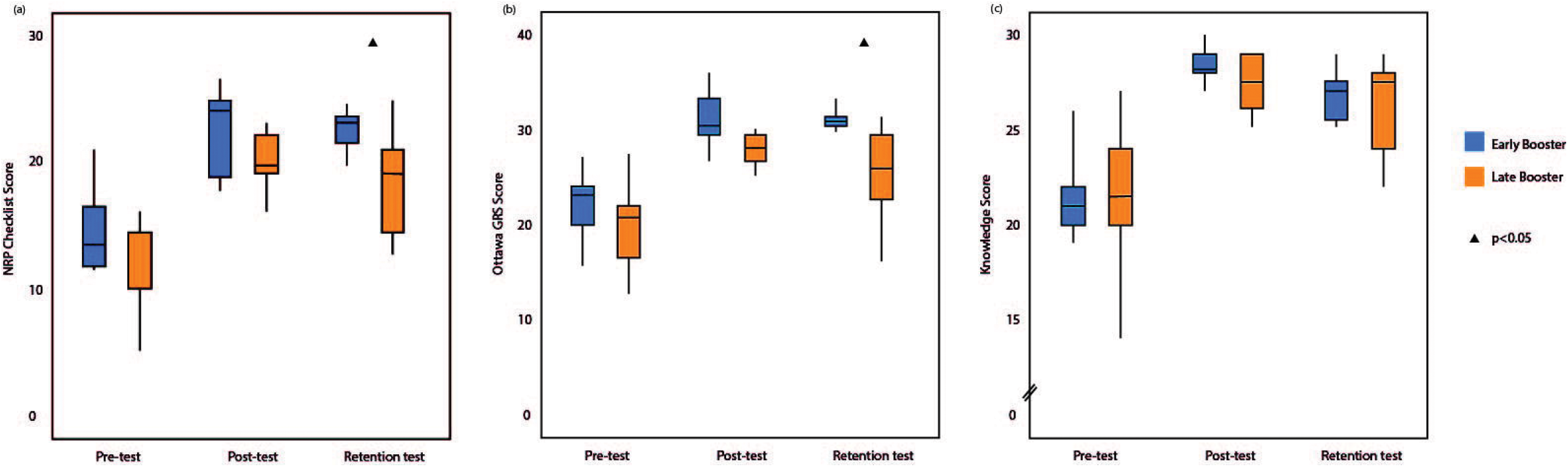
Boxplots of NRP checklist, Ottawa GRS and knowledge test scores at each experimental stage.

## Discussion

In this study, we found that an early refresher after NRP training was more effective at promoting long-term retention of technical and non-technical skills compared to a late refresher, but has no impact on knowledge retention.

We have shown that a quantitative theoretical framework underpinning retention in declarative memory can be translated to neonatal resuscitation – a high-stakes skill set that requires mixed declarative and procedural memory, executed under intense time and situational pressure. It is not enough that learning is distributed, as discovered by Kaczorowski et al^9^; instead, the 10-30% interval rule for ISI:RI ratios ^25^ also applies to complex, mixed-memory skills such as NRP.

We have also shown that an early refresher improves retention of non-technical skills. It has been shown previously that technical and non-technical skills are correlated, and that residents who are proficient in one domain also demonstrate proficiency in the other^35^. This group postulated that the reason may be due to the concept of ‘cognitive load’. Working memory is limited, whilst long-term memory has much greater capacity. Participants who are technically skilled have committed these aspects to long-term memory, and are therefore able to free cognitive resources to better confront non-technical challenges.

We found no evidence for improved knowledge retention in the early refresher group. This is perhaps surprising, given that multiple choice questions are the closest test of purely declarative memory. It is notable that the knowledge exam was both the highest scoring of the three outcome measures, and the one to suffer the least attrition from immediate post-test to retention. We speculate that the cognitive demands were too low to show an effect from the intervention, or that a ceiling effect applies whereby the benefit of the early refresher is only apparent when attrition of memory reaches a certain threshold.

Health systems invest a large amount of time and money so that professionals respond appropriately in resuscitation situations. One trial estimated that the cost of an adult cardiopulmonary resuscitation program per life saved was around $500,000 and that a significant proportion of this was due to training^36^. For neonatal resuscitation, the evidence suggests that this investment in time and money does not lead to the retention of skills for a clinically desirable time period^9,37^. At present, there is no consensus about the optimal way to structure training for high-stakes scenarios such as resuscitation and this is a recurrent focus of gaps-in-knowledge summaries from ILCOR^2,12^. Most authorities currently take an interval-based approach to training and certification, but with varying time-frames. In North America, it is recommended that trainees complete the program every two years, whilst in the UK certification is valid for four. There is no nationally-mandated training to be completed in between these intervals. This heterogeneity in practice, and the paucity of the evidence base, is a major priority for future research efforts.

Our work suggests a way forward based on an evidence-based theoretical framework. Policy makers must first decide what time-period retention of performance is to be maintained over – this will depend on an analysis of service needs and cost-benefit ratio – and then structure a training program in such a way that refresher sessions are delivered at appropriately-distributed intervals. We used a retention interval of four months for the purposes of this study. In reality, healthcare systems are likely to desire longer recertification cycles. Future studies should aim to determine optimal refresher timings and retention intervals for real-world use, and to investigate alternative methods of delivering distributed learning (e.g. as online courses or as other forms of distance learning).

Although we studied neonatal resuscitation, these findings may also be generalizable to other resuscitation programs such as Adult Cardiac Life Support, and to other interventions where skill retention depends on mixed types of memory.

There are limitations to our study. We were unable to recruit as many participants as we had intended, and the study is therefore underpowered. However, as there was a significant difference in our primary outcome, the issue of type 2 error is minimal. We plan to address this with a larger study in future, and are considering strategies that will mitigate these problems – for instance, a multi-centre trial that will both increase both generalizability and the target recruitment pool. We studied performance in a simulated situation, which is common for resuscitation studies because of the difficulties in controlling for variables in real resuscitation scenarios. There is good evidence to suggest that performance in a simulated setting can be transferable to clinical practice^38^.

Ultimately, the aim of such research should not just be about improving education, but about improving patient outcome. There is evidence that neonatal resuscitation training improves APGAR scores in the US^8^, decreases the incidence of meconium aspiration in France^39^, shortens the duration of hospitalization in Turkey^40^, and reduces neonatal mortality in China^6^. There is scope for future studies to build on our work, and look at changes in patient outcome following introduction of a teaching program guided by our conclusions.

## Data Availability

Please email corresponding author for data files.

## Acknowledgements

We would like to thank Doaa Gazarin, Haytham Qosa, Emily Hladkowicz, Sam Caldbick, Diana Noseworthy, Meredith Mackay, Alvi Rahman, Karl Schebesta, Mounir Fayez, and Dan Power for acting as confederates during the simulation scenarios.

## Appendix 1

Neonatal Resuscitation Program Megacode Assessment – Canadian Adaptation

| **Item** | **Points** |
| --- | --- |
| Checks Equipment inc. Bag, Mask & Oxygen Supply | 2 |
| Asks 3 assessment questions (term, tone, crying or breathing) | 2 |
| Positions head, clears airway if necessary | 2 |
| Dries, removes wet towels, repositions head | 2 |
| Requests description of respirations and heart rate | 2 |
| Indicates need for positive pressure ventilation | 2 |
| Applies pulse oximeter appropriately | 2 |
| Provides positive pressure ventilation correctly | 2 |
| Checks for rising heart rate, breath sounds within 5-10 seconds | 2 |
| Takes corrective action when not improving | 2 |
| Administers oxygen to meet targeted saturations using pulse oximeter and oxygen blender | 2 |
| Re-evaluates heart rate | 2 |
| Identifies need to start chest compressions | 2 |
| Demonstrates correct compression technique | 2 |
| Demonstrates correct rate and co-ordination with ventilation | 2 |

## Appendix 2

### Neonatal Resuscitation Knowledge Test

#### Instruction to candidates

For each question stem, **circle the <u>single</u> best answer** from the options provided.

#### Questions

1. Approximately what percentage of newborns will require some assistance to begin regular breathing?
  a. 1%
  b. 5%
  c. 10%
2. Approximately what percentage of newborns will require extensive resuscitation to survive?
  a. 1%
  b. 5%
  c. 7%
3. The air that fills the baby’s alveoli during normal transition contains:
  a. 21% oxygen
  b. 40% oxygen
  c. 100% oxygen
4. If a baby does not begin breathing in response to stimulation, you should:
  a. Provide positive pressure ventilation
  b. Intubate the trachea
  c. Perform tracheal suctioning for meconium.
5. Restoration of adequate ventilation will usually improve the baby’s heart rate
  a. Rapidly
  b. Gradually
  c. Slowly
6. True or false: *A vigorous term newborn with meconium in the amniotic fluid will need to have his trachea suctioned via an endotracheal tube?*
  a. True
  b. False
7. When deciding which babies need tracheal suctioning, the term ‘vigorous’ is defined by what 3 characteristics?
  a. Strong respiratory efforts, good muscle tone, heart rate greater than 100 beats per minute.
  b. Strong respiratory efforts, good muscle tone, good skin colour.
  c. Strong respiratory efforts, good skin colour, heart rate greater than 100 beats per minute.
8. When suctioning a newborn’s nose and mouth with a bulb syringe or suction catheter, the correct order is:
  a. Mouth, then nose.
  b. Nose, then mouth.
9. Once resuscitation has been initiated, how often is it recommended that the team re-evaluate the baby’s condition?
  a. 45 seconds
  b. 30 seconds
  c. 60 seconds
10. Which of the following is a recommended method of stimulating an apneic newborn?
  a. Slap the back
  b. Squeeze the rib cage
  c. Flick the sole of the foot
11. True or false: *In the delivery room, the oximetry probe should always be placed on the baby’s right hand or wrist*.
  a. True
  b. False
12. True or false: *Oxygen saturation should be expected to be >85% by 2 minutes of age*.
  a. True
  b. False
13. You have stimulated a newborn and suctioned her mouth. It is now 30 seconds after birth, and she is still apneic. Her heart rate is 80 beats per minute. Your next action should be:
  a. Continue stimulation, and give free-flow supplemental oxygen.
  b. Provide positive pressure ventilation.
  c. Begin chest compressions.
14. The color of which body parts should be used to assess the baby’s state of oxygenation?
  a. Lips, tongue and torso
  b. Hands and feet
15. Resuscitation of the term newborn should begin with what percentage of oxygen?
  a. 21%
  b. 40%
  c. 60%
16. When ventilating a newborn, you should provide positive-pressure ventilation at a rate of:
  a. 20 – 40 breaths per minute
  b. 30 – 40 breaths per minute
  c. 40 – 60 breaths per minute
17. What initial inspiratory pressure should you aim to begin positive pressure ventilation with?
  a. 10 cmH_2_O
  b. 20 cmH_2_O
  c. 30 cmH_2_O
18. The ‘S’ in MR SOPA stands for:
  a. Start chest compressions
  b. Suction mouth and nose
  c. Saturation probe
19. The ‘O’ in MR SOPA stands for:
  a. Open mouth
  b. Oxygen
  c. Orogastric tube
20. When providing positive-pressure ventilation with a mask for more than a few minutes, what device should be inserted to vent gas from the stomach?
  a. Endotracheal tube
  b. Meconium aspirator
  c. Orogastric tube
21. You have provided effective positive-pressure ventilation for 30 seconds and the baby’s heart rate is below 60 beats per minute. Your next action should be:
  a. Reposition the head
  b. Insert an endotracheal tube
  c. Start chest compressions
22. The baby is now breathing and its heart rate is >100 bpm. It is now safe to discontinue positive-pressure ventilation.
  a. True
  b. False
23. The preferred method of delivering chest compressions is the:
  a. Thumb technique
  b. 2-finger technique
  c. Palm technique
24. The correct depth of chest compressions is about:
  a. One-quarter the anterior-posterior diameter of the chest
  b. One-third the anterior-posterior diameter of the chest
  c. One-half the anterior-posterior diameter of the chest
25. During positive-pressure ventilation with chest compressions, the rate of total events per minute (compressions plus breaths) should be:
  a. 60 events per minute
  b. 100 events per minute
  c. 120 events per minute
26. After 30 seconds of chest compressions, you stop and count 8 heartbeats in 6 seconds. What is the baby’s heart rate?
  a. 48 beats per minute
  b. 60 beats per minute
  c. 80 beats per minute
27. You have performed the corrective sequence for inadequate positive-pressure ventilation (MR SOPA). The baby’s heart rate fails to rise and bilateral breath sounds are not heard while you are using a bag and mask. Which of the following actions is most appropriate?
  a. Insert an umbilical vein catheter and give IV epinephrine
  b. Insert a laryngeal mask airway or an endotracheal tube
  c. Increase the rate of positive-pressure ventilation
28. When using an anatomically shaped mask, which end should be placed over the infant’s nose?
  a. Pointed
  b. Rounded
29. When positioning an infant’s head for positive-pressure ventilation, which position is appropriate?
  a. Flexed
  b. Extended
  c. Sniffing
30. What 3 things must be determined first during your initial neonatal resuscitation assessment?
  a. Gestational age, respiratory effort or crying, muscle tone
  b. Muscle tone, presence of meconium, respiratory effort or crying
  c. Heart rate, gestational age, skin colour

